# Sequential Deep Learning to Predict Non-Central to Central Geographic Atrophy Progression from OCT Imaging

**DOI:** 10.64898/2026.06.17.26355882

**Authors:** Sadia Siraz, Hindolo Kamanda, Ahammed Sakir Nabil, Sina Gholami, Nitya Tangada Rao, Sally S. Ong, Minhaj Nur Alam

**Author notes:** **Corresponding Author:** Minhaj Nur Alam, Ph.D, 9201 University City Boulevard, Charlotte, NC 28223. Co Corresponding Author: Sally S. Ong, M.D, Wake Forest University School of Medicine, Winston-Salem, NC, USA. **Meeting Presentation** Previously presented on SPIE Photonics West 2026 (San Francisco, CA; January 17-22, 2026); Will be presented at ARVO Annual Meeting 2026 (Denver, Colorado; May 3-7, 2026).

## Abstract

**Purpose:** To develop and validate a temporal deep learning framework for predicting geographic atrophy (GA) progression across multi-year horizons using longitudinal optical coherence tomography (OCT) sequences.

**Design:** Retrospective longitudinal cohort study.

**Subjects, Participants, and/or Controls:** A total of 91 patients with dry age-related macular degeneration (AMD) were identified from Wake Forest University School of Medicine (2013-2023), yielding 455 OCT volumes. Two prediction cohorts were defined: 32 patients with no GA (NGA) at baseline who subsequently developed GA, and 35 patients whose earliest GA manifestation was non-central GA (NCGA). Non-progressing patients served as negative controls.

**Methods:** OCT B-scan volumes were encoded into visit-level feature representations using three pretrained architectures (ResNet-18, ResNet-50, ViT-B/16). Chronologically ordered visit embeddings, optionally augmented with inter-visit time intervals (Δt), were processed through recurrent neural networks (RNN), long short-term memory networks (LSTM), and Transformer encoders to model longitudinal disease trajectories. Models were trained and evaluated independently for prediction horizons of 2, 3, 4, 5, and 6 years using patient-level stratified splits (80/20). Performance was assessed across five random seeds.

**Main Outcome Measures:** Area under the receiver operating characteristic curve (ROC-AUC), F1-score, and accuracy for predicting two clinically critical transitions: NGA to GA onset and NCGA to central GA (CGA) involvement.

**Results:** For NGA to GA prediction, models achieved ROC-AUC of 0.84-0.94 at 2-4 years and 1.00 at 5-6 years. For NCGA to CGA prediction, Transformer-based models achieved peak AUC of 0.95 (±0.11) at 4 years and 0.96 (±0.06) at 5 years. Longer input sequences (8 visits vs. 4 visits) consistently improved NCGA to CGA performance at extended horizons. Temporal interval encoding improved stability in several LSTM configurations.

**Conclusions:** Temporal deep learning applied to longitudinal OCT sequences can predict GA progression across clinically meaningful 2-6 year horizons without pixel-level annotations. These findings support the feasibility of automated, individualized risk stratification to guide complement inhibitor therapy decisions in patients with GA.

## Introduction

Age-related macular degeneration (AMD) represents one of the primary causes of permanent vision loss worldwide, impacting roughly 20% of individuals over 65, with projected global cases reaching 288 million by 2040.^1^ The disease presents in two forms: dry or non-neovascular AMD (approximately 80% of cases) and neovascular AMD.^2^ Geographic atrophy (GA) is the late stage of dry AMD, involving progressive loss of retinal pigment epithelium (RPE), photoreceptors, and underlying choriocapillaris,^5^ affecting approximately 160,000 new U.S. patients annually and responsible for roughly 20% of AMD-related legal blindness.^6,7^ GA typically begins peripherally (non-central GA; NCGA) before advancing toward the fovea over multiple years ^8^. Once atrophy involves the fovea (central GA; CGA),^9^ patients experience devastating functional impairment affecting reading, driving, and facial recognition with 57% of NCGA eyes progressing to CGA within 4 years.^9,10^ While anti-vascular endothelial growth factor (VEGF) treatments have revolutionized neovascular AMD management^3^ (inhibiting growth of neovascularization), therapeutic options for GA remained unavailable until 2023.^4^ With the 2023 FDA approval of pegcetacoplan and avacincaptad pegol, the first complement inhibitors shown to slow growth rate of GA lesions,^11^ the clinical need for predicting GA progression has taken center stage. Pegcetacoplan achieved approximately 22-26% reduction in GA growth rate overall compared to sham at 24 months in the OAKS/DERBY trials,^12^ with extension data from GALE confirming increasing efficacy over 36 months and particularly pronounced benefit in extrafoveal lesions (up to 42% reduction vs. projected sham).^13^ A robust prognostic model can help prioritize which patients would benefit most from currently available treatments, which slow GA progression but do not reverse atrophy.

Optical coherence tomography (OCT) is one of the most widely used imaging modalities for longitudinal assessment of progression of retinal pathologies. While fundus autofluorescence (FAF) remains standard for GA area quantification in clinical trials,^14^ OCT’s volumetric, depth-resolved imaging enables superior detection of temporal changes in RPE integrity, photoreceptor structure, and choroidal hypertransmission which are critical biomarkers preceding complete RPE and outer retinal atrophy.^15,16^ OCT distinguishes incomplete versus complete outer retinal atrophy (iRORA/cRORA), precisely measures the distance between atrophy to the fovea, and detects nascent changes months before disease manifestation on FAF.^17,18^ The Classification of Atrophy Meetings (CAM) consensus established validated OCT-based GA and atrophy definitions,^18^ and OCT’s widespread availability with standardized acquisition protocols positions it ideally for temporal progression modeling.^19^

Despite robust anatomical biomarkers, predicting individual trajectories remains exceptionally challenging. GA growth rates vary dramatically, influenced by lesion morphology, drusen characteristics, genetics, and fellow-eye status.^10,20^ Pre-atrophic features such as hyperreflective foci, reticular pseudodrusen, and hypertransmission defects show complex, non-linear relationships to future atrophy development and foveal involvement.^21–23^ Human assessment of progression risk across hundreds of B-scans and multiple timepoints exceeds practical clinical capacity and introduces substantial variability. Clinicians lack validated tools for quantitative, patient-specific risk stratification, particularly for identifying which NCGA eyes face imminent foveal involvement. Combined with an aging population and rapidly increasing quantities of longitudinal imaging, manual OCT review for thousands of at-risk patients is unsustainable. Deep learning systems offer a scalable solution, extracting subtle spatiotemporal patterns invisible to humans while providing objective risk scores and enabling personalized surveillance strategies.^24,25^

Recent Artificial intelligence (AI) advances in AMD have emphasized GA detection and short-term growth prediction rather than long-term changes. Deep learning models have predicted GA area expansion from baseline FAF or OCT and generated individualized future GA lesion maps from single-visit volumes.^25–27^ Several approaches have targeted intermediate AMD to GA conversion or early atrophy development, including short-term OCT-based prediction (DeepGAze),^28^ machine-learning identification of OCT biomarkers predictive of atrophy, and deep survival models leveraging longitudinal OCT volumes.^29^ Complementary work has focused on detecting nascent GA and related pre-atrophic changes using OCT-based deep learning,^30,31^ and multimodal frameworks have highlighted the prognostic value of CAM-defined features and multimodal imaging signatures.^24,25^

Beyond GA-specific applications, sequence-based architectures such as convolutional neural network (CNN) - long short term memory (LSTMs), time-aware LSTMs, and Transformers have improved late AMD progression prediction from longitudinal fundus photographs and OCT.^32–35^ Vision Transformers and modern CNN backbones such as ResNets have become standard for retinal imaging tasks, including AMD classification from both fundus photographs and OCT.^36–38^ However, existing models typically address GA detection, binary onset prediction, single-visit imaging, or short prediction horizons. *<u>Critically, no study has modeled longitudinal OCT sequences to predict multi-year NGA to GA</u> <u>and NCGA to CGA progression, which is clinically essential given location-dependent therapeutic</u> <u>efficacy of complement inhibitors</u>*.^39–41^ This gap persists because: (1) most prior work employed single-timepoint imaging or follow-up periods insufficient for multi-year modeling^25–27,29^, (2) longitudinal OCT datasets with extended GA outcomes remain scarce;^29^ (3) temporal architectures (RNNs, LSTMs, Transformers) have not been systematically applied to OCT-based GA progression despite their success in other AMD progression tasks;^32–35^ and (4) the clinical imperative to forecast NGA to GA and NCGA to CGA progression emerged only recently with complement inhibition therapies that can slow GA growth towards the fovea and prolong preservation of central vision.^40,41^

We previously reported deep learning methods for GA classification using OCT from the same patient population.^42^ However, classification of existing GA does not address the prognostic challenges surrounding multi-year disease progression, motivating the present extension to longitudinal modeling. In this study, we have developed a comprehensive temporal deep learning framework for multi-horizon GA progression prediction using longitudinal OCTs. Our contributions include:

1. A deep-learning-based prognosis model capable of predicting NGA to GA and NCGA to CGA progression using visit-level labels rather than pixel-level annotations;
2. Model validation using a private clinical dataset of OCT scans from 91 patients from the Wake Forest University School of Medicine;
3. A systematic comparison of different temporal modeling strategies to address variability in patient follow-up patterns; and
4. Generation of 2-6 year progression risk estimates, a clinically crucial window for complement inhibitor therapy where timing is essential to prevent irreversible foveal involvement.

## Methodology

### Dataset

#### Study Design and Data Source

We conducted a retrospective longitudinal cohort study utilizing clinical data from Wake Forest University School of Medicine over a decade-long period from January 1, 2013, to January 28, 2023. The study protocol received institutional review board (IRB) approval and adhered to the tenets of the Declaration of Helsinki. Patients were systematically identified through ICD-9 codes 362.50 and 362.51 and ICD-10 codes H35.30 and H35.31.

Patients were included if they had longitudinal OCT imaging with adequate follow-up to assess GA progression. Exclusion criteria included: dry AMD without evidence of GA development; subfoveal GA at baseline; absence or non-retrievability of longitudinal OCT imaging; fewer than two available OCT visits; coexisting ocular pathologies likely to confound OCT interpretation including neovascular AMD, diabetic retinopathy, glaucoma, retinal vascular occlusions, retinal detachment, and macular dystrophies; and prior retinal treatments. After applying these criteria, 91 patients with high-quality imaging were retained for analysis. Each patient contributed a single predefined study eye (OD 48.08%, OS 51.92%) that remained consistent across all visits. The dataset encompassed 455 OCT volumes (a total of 11513 B-Scans), comprising 258 volumes labeled as CGA (6522 B-Scans), 74 as NCGA (1874 B-Scans), and 123 as NGA (3117 B-Scans). Representative OCT B-Scans are shown in figure 2b.

#### Cohort Characteristics and Image Acquisition

All OCT volumes were acquired using a standardized protocol on the Heidelberg HRA+OCT Spectralis platform ^43^ with Heidelberg Heyex software. Most OCT volumes contained 25 B-scans (512×496×25 voxels) reflecting the standard acquisition protocol. In the CGA cohort, 242 volumes contained 25 B-scans, while 14 and 2 volumes contained 31 and 19 B-scans, respectively. Similarly, the NCGA and NGA cohorts each exhibited minor deviations, with 4 and 7 volumes containing 31 B-scans, respectively.

All OCT volumes underwent systematic evaluation by a board-certified retinal specialist (SSO) and an ophthalmologist in subspecialty training to become a retinal specialist (NTR). GA classification followed established minimum criteria for incomplete RPE and outer retinal atrophy (iRORA) as defined by the Classification of Atrophy Meetings (CAM): loss of the outer retina, loss of the retinal pigment epithelium, and choroidal hypertransmission with no requirement for the lesion to be at least 250um in size. NGA was defined as the absence of GA features, NCGA as GA localized exclusively outside the central 1 mm ETDRS circle, and CGA as GA extending into or originating within the central 1 mm ETDRS circle.^44^ Two experienced graders independently assigned ordinal disease severity grades (scale: 1–3; 1 = NGA, 2 = NCGA, 3 = CGA) at each patient visit. Inter-rater reliability was evaluated at the visit level using percent agreement and Cohen’s kappa (κ), with quadratic-weighted kappa calculated to account for the ordered nature of the grading scale.

#### Longitudinal Progression and Dataset Composition

Patients demonstrated variable visit frequencies and follow-up intervals, capturing diverse progression trajectories including NGA to NCGA to CGA, direct NGA to CGA transitions, NCGA to CGA progression, and stable non-progressors. For the NGA to GA prediction task, we selected 32 patients who exhibited no GA at baseline and subsequently developed GA during follow-up. For the NCGA to CGA prediction task, we constructed a cohort of 35 patients who initially presented with NCGA at baseline or who began as NGA but converted to NCGA and subsequently CGA. Both modeling cohorts were defined by disease stage-specific baselines rather than unique patient identifiers, allowing individuals to contribute to multiple progression pathways if they experienced sequential transitions. Specifically, 5 patients who experienced sequential NGA to NCGA to CGA transitions were included in both cohorts - for the NGA to GA cohort, their data was indexed from their first NGA visit, whereas for the NCGA to CGA cohort, their data was independently re-indexed from the first visit at which NCGA was documented. By re-indexing patients at the time they met new disease stage criteria, we ensured prognostic modeling was anchored to the appropriate disease stage without information leakage. The index visit served as the starting point for prediction and was defined as the first visit at which the patient met the baseline disease stage relevant to the task. The patient selection process is illustrated in Figure 1

**Figure 1:**
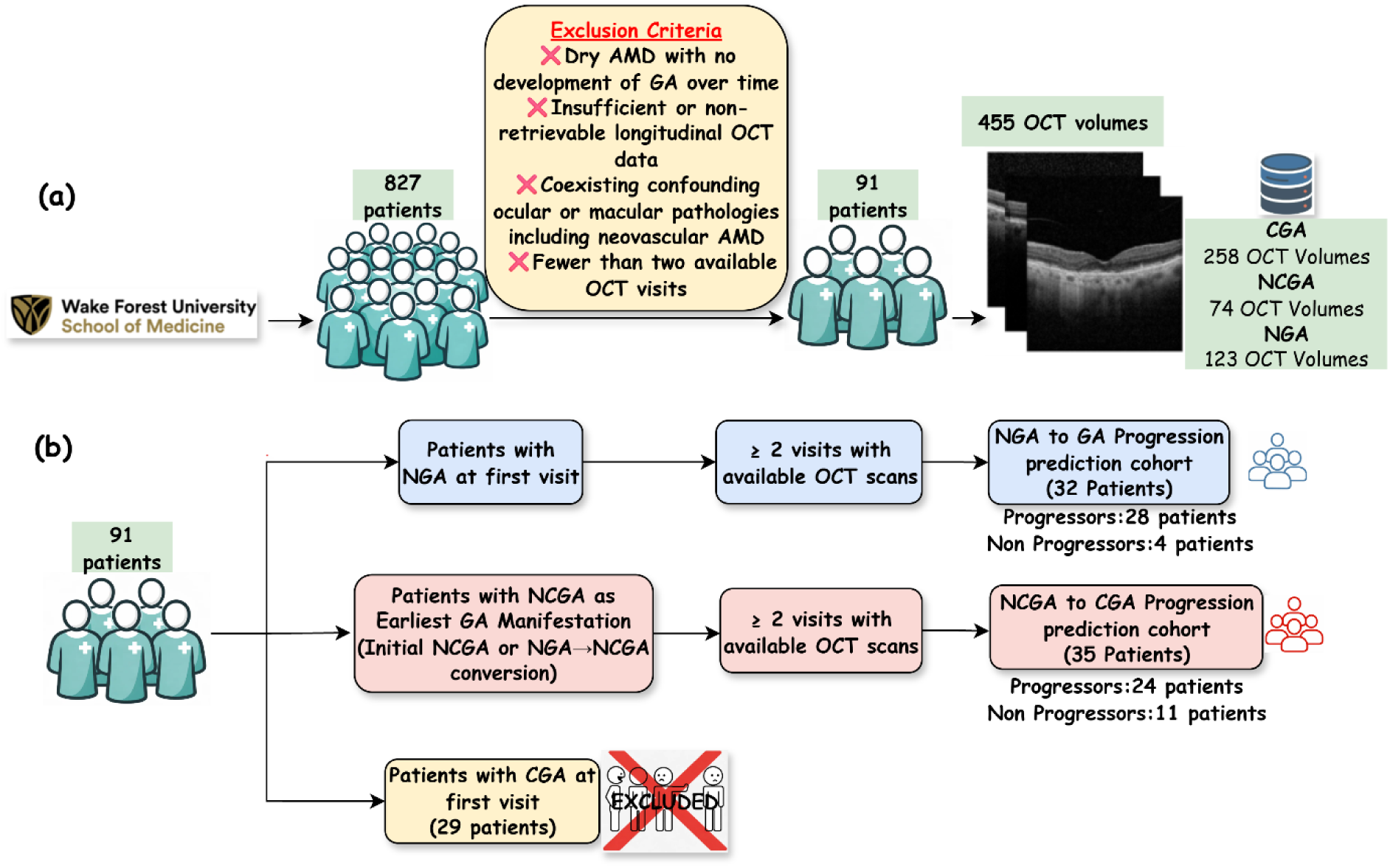
Study cohort selection and construction of longitudinal GA progression prediction cohorts from OCT imaging. (a) Dataset assembly and filtering process; (b) Construction of prediction cohorts based on baseline disease stage.

Within the NGA cohort (n = 32), 28 patients (87.5%) developed GA during follow-up with a mean time to conversion of 3.0 years (range: 2 months - 7.9 years). Among the NCGA cohort (n = 35), 24 patients (68.6%) progressed to central involvement with a mean time to conversion of 2.6 years (range: 4.1 months - 6.6 years). Non-progressing patients served as negative controls. The progression cohort characteristics can be found in figure 2a.

**Figure 2.**
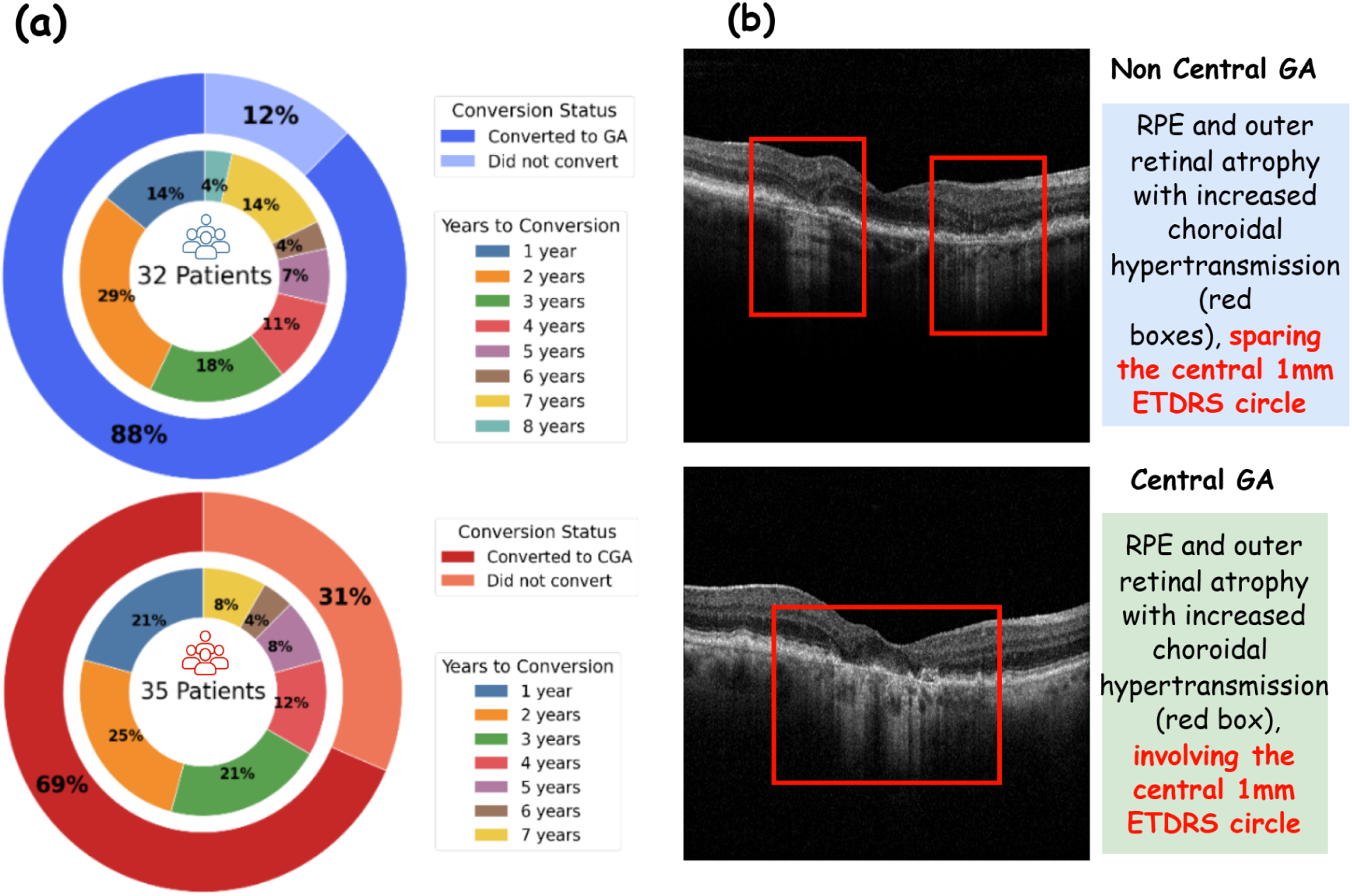
Progression cohort characteristics and representative OCT features of NCGA and CGA. (a) Distribution of patients and time-to-conversion for the two progression tasks: NGA to GA (n = 32) and NCGA to CGA (n = 35). Outer rings indicate the proportion of progressors and non-progressors, while inner segments illustrate the number of years to disease conversion. (b) Representative OCT B-scan examples illustrating structural differences between NCGA and CGA.

Patient visit contributions ranged from 2 to 26 examinations, with longitudinal follow-up extending to more than 10 years. Prediction models were developed for multiple clinically relevant time horizons (i.e., the future follow-up period over which disease progression was predicted) of 2, 3, 4, 5, and 6 years. The selection of maximum sequence lengths for deep learning models (7 and 19 visits for NGA to GA progression; 4 and 8 visits for NCGA to CGA progression) was empirically determined based on visit frequency distributions, balancing data utilization with computational efficiency.

For each prediction task and time horizon, patients were re-indexed at the first visit meeting the task-specific baseline disease stage. Prediction labels were defined relative to this index visit, with patients included only if sufficient follow-up was available to determine progression status within the specified horizon. This horizon-specific cohort construction and indexing strategy is formally described in Algorithm 2.

### Data Preprocessing

All OCT B-scans were resized to 224×224 pixels and converted to 3-channel RGB format. Pixel intensities were normalized using ImageNet statistics (mean = [0.485, 0.456, 0.406], standard deviation = [0.229, 0.224, 0.225]) for ResNet models and standardized normalization (mean = 0.5, standard deviation = 0.5) for Vision Transformer (ViT-B/16).

### Feature Engineering and Dataset Assembly

#### Feature Extraction

Our pipeline does not rely on lesion annotation or segmentation. Visit-level feature representations were extracted using three ImageNet-pretrained architectures as shown in figure 3:

- ResNet-18: 512-dimensional feature vectors
- ResNet-50: 2048-dimensional feature embeddings
- Vision Transformer (ViT-B/16): 768-dimensional feature representations

For each prediction horizon and sequence length, OCT features were re-extracted from the specific visits corresponding to that configuration, avoiding reuse of future or out-of-window scans. Feature vectors from all constituent B-scans per visit were aggregated using mean pooling to generate a single unified representation. Padded visits utilized zero vectors to maintain consistent tensor shapes. This visit-level representation learning process, including chronological ordering of visits, mean pooling of B-scan features within each visit, and encoder-specific feature dimensionality, is summarized in Algorithm 1.

##### Algorithm 1

Visit-level Representation Learning from Longitudinal OCT

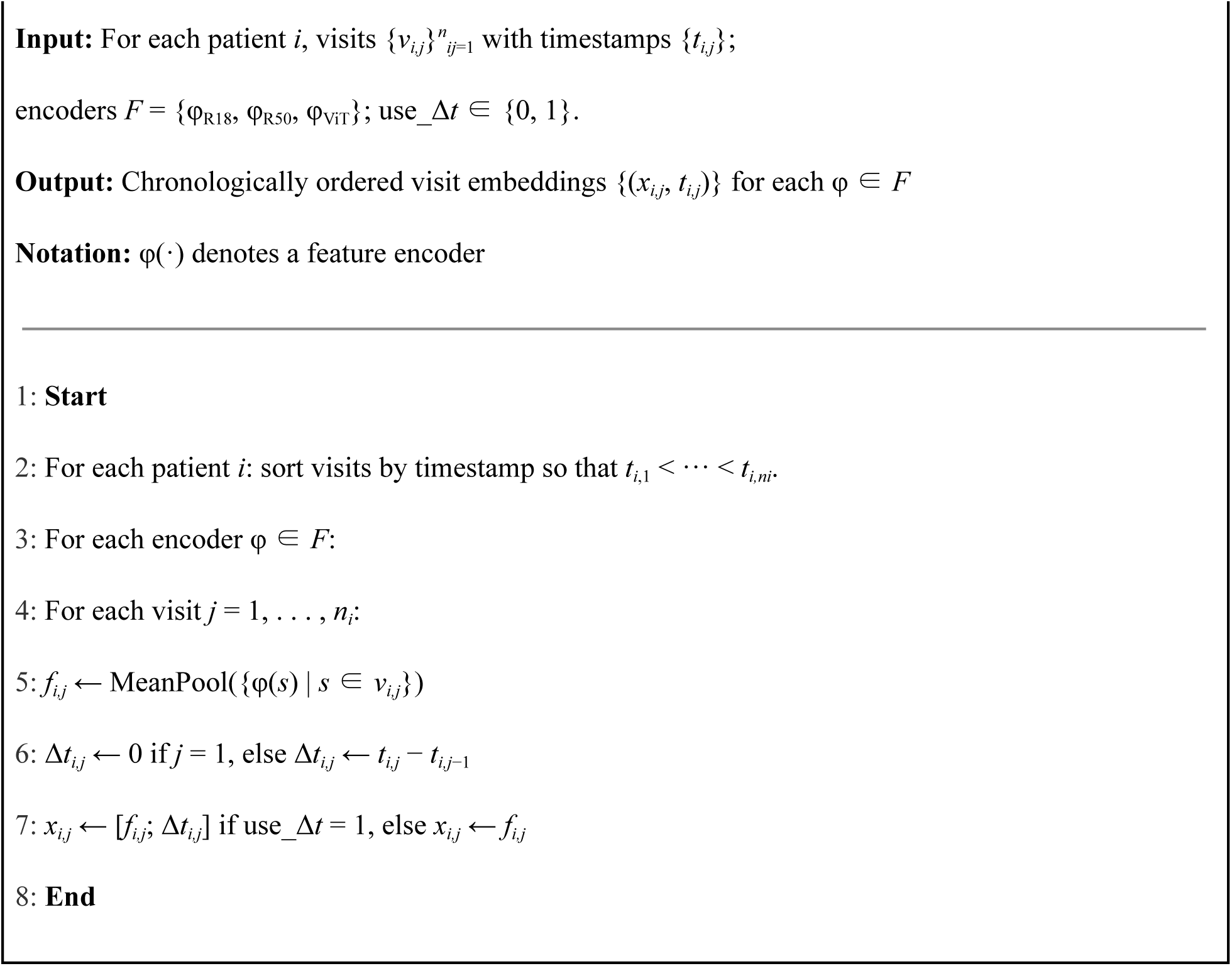

For patient *i*, the *j*-th clinical visit is denoted as *v_i_, □* and acquired at timestamp *t_i_, □*, with visits ordered chronologically such that *t*_*i*,1_ < ··· < *t*_*i,ni*_. Each visit *v_i_, □* consists of multiple OCT B-scans, from which a visit-level image representation *f_i,j_* is obtained by mean pooling deep features extracted from all constituent B-scans using a pretrained encoder. To incorporate longitudinal timing information, the elapsed time between consecutive visits is defined as Δ*t_i,j_* = *t_i,j_* − *t_i,j_*_−1_ for j > 1, with *Δt_i_,*₁ = 0. When temporal augmentation is enabled, *Δt_i_, □* is concatenated to the image embedding to form the final visit representation *x_i_, □* = [*f_i_, □; Δt_i_, □*]; otherwise, *x_i_, □ = f_i_, □*.

**Figure 3:**
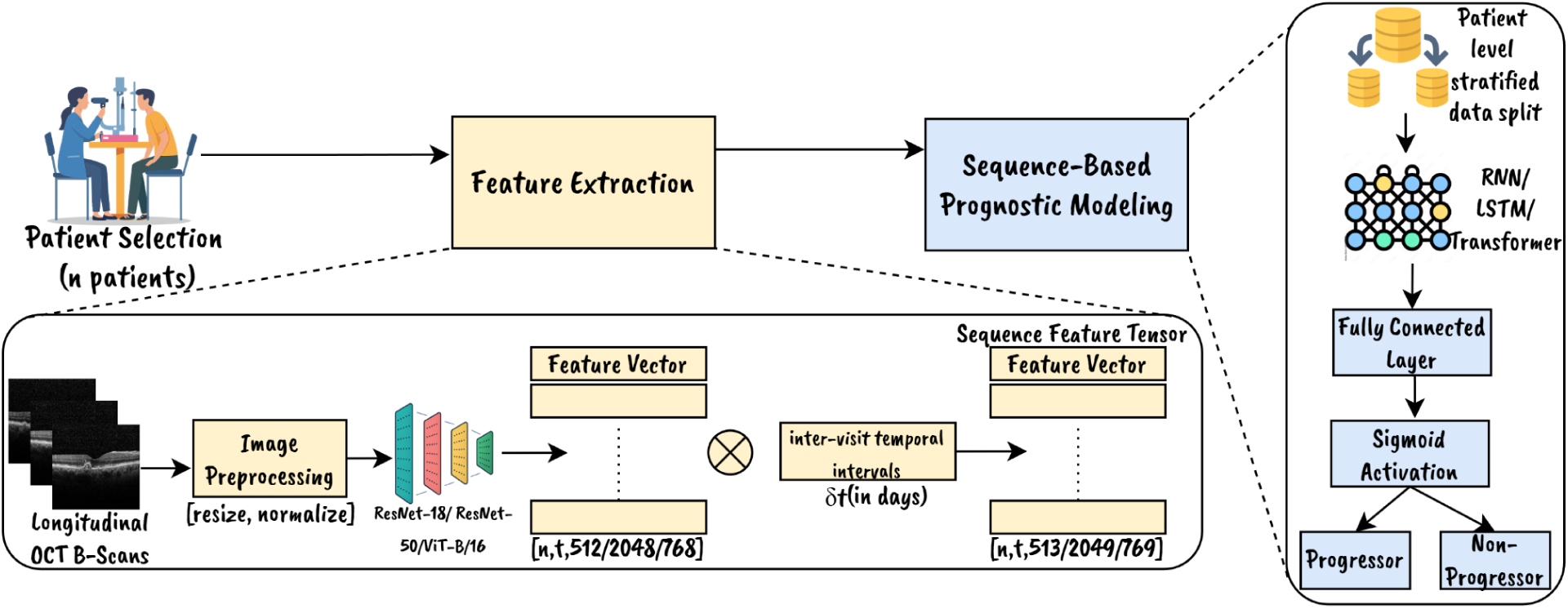
Overview of the longitudinal GA progression study design and modeling framework.

#### Temporal Encoding

To capture longitudinal disease dynamics, temporal information was incorporated through delta time (Δt) augmentation. The elapsed time between consecutive visits (in days) was concatenated to each visit embedding, resulting in augmented feature vectors with +1 dimension. Both temporally augmented and standard feature configurations were evaluated. When enabled, temporal augmentation was implemented by concatenating the elapsed time between consecutive visits (Δt) to each visit embedding, as detailed in Algorithm 1.

#### Dataset Assembly, Sequence Processing, and Model Input

Patient longitudinal trajectories were structured as tensors with shape *(num_visits× feature_dim)*, paired with corresponding visit-level progression labels. Each patient visit was represented as a feature vector extracted from OCT B-Scans using the ImageNet-pretrained architectures. *Δt* was concatenated to each vector when temporal augmentation was used.

Variable-length sequences were truncated to retain the most recent visits up to the prediction index time and zero-padded to a fixed maximum sequence length using dummy visits with null feature vectors, zero temporal intervals, and sentinel labels (label = −1). Binary attention masks, constructed based on the number of retained visits per patient sequence, excluded padded entries during model training and evaluation. Models received chronologically ordered sequences with attention masks applied to ignore padded entries, producing a single probability score corresponding to the last valid (most recent) visit in each patient sequence. Progression labels were aligned with the final valid visit of each patient sequence for all prediction time horizons. Sequence truncation to the most recent visits, zero-padding to fixed lengths, construction of binary validity masks, and alignment of labels with the last valid visit are operationalized as outlined in Algorithm 2.

For each prediction task and horizon *h* ∈ *{2, 3, 4, 5, 6}* years, patient sequences were indexed at the first visit meeting the task-specific baseline disease stage, referred to as the index visit. Let *X_i_*⁽*ᴸ*⁾ ∈ ℝ*ᴸˣᵈ* denote the longitudinal feature sequence for patient *i*, constructed by retaining the *L* most recent visits prior to and including the index visit and zero-padding when fewer than *L* visits were available. A corresponding binary mask *M_i_*⁽*ᴸ*⁾ ∈ *{0,1}ᴸ* indicates valid (non-padded) visits in the sequence. All temporal models process chronologically ordered sequences and generate predictions based on the hidden representation associated with the last valid visit, as identified by *M_i_*⁽*ᴸ*⁾.

##### Algorithm 2

Longitudinal Modeling for GA Progression Prediction

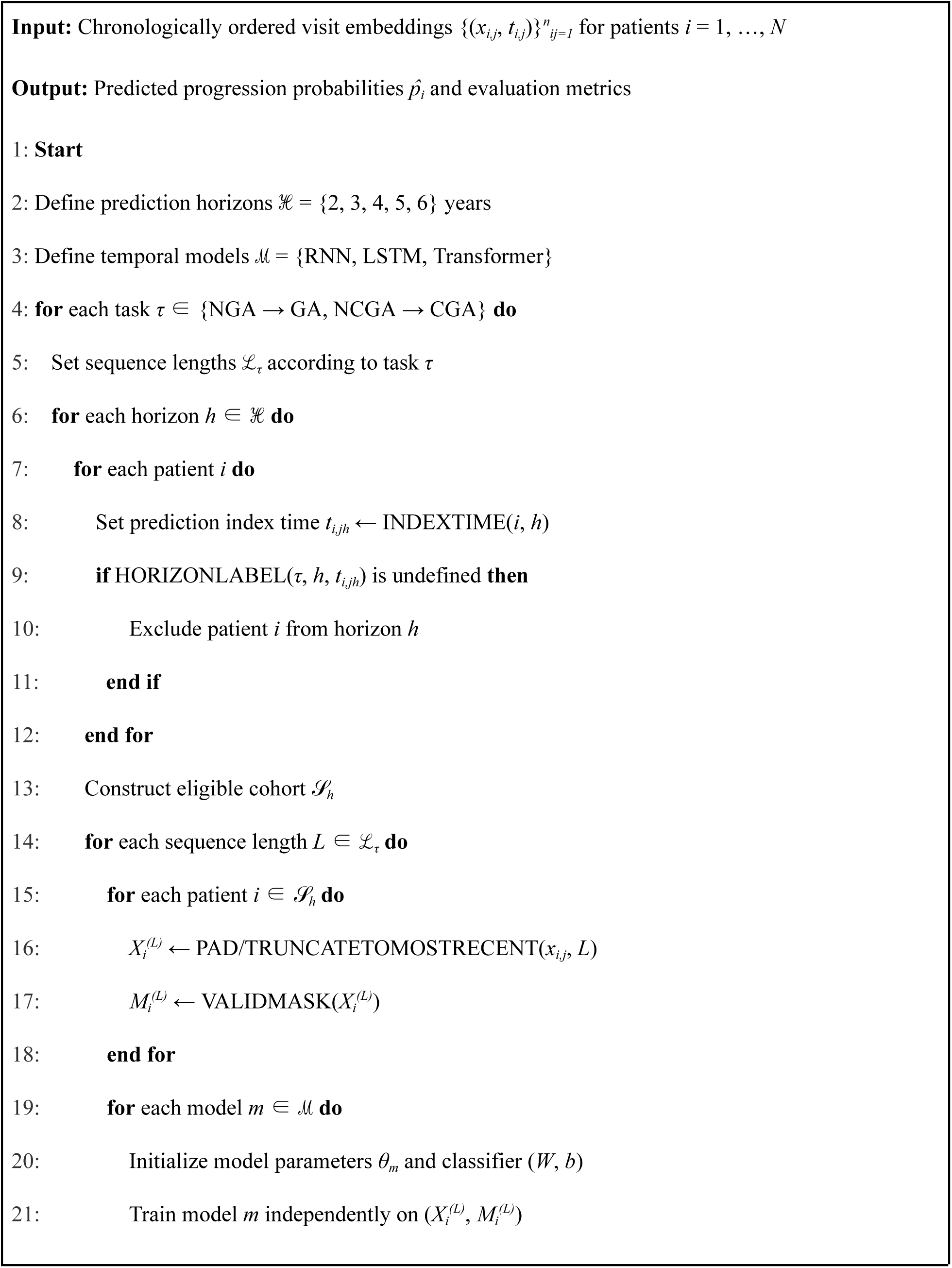
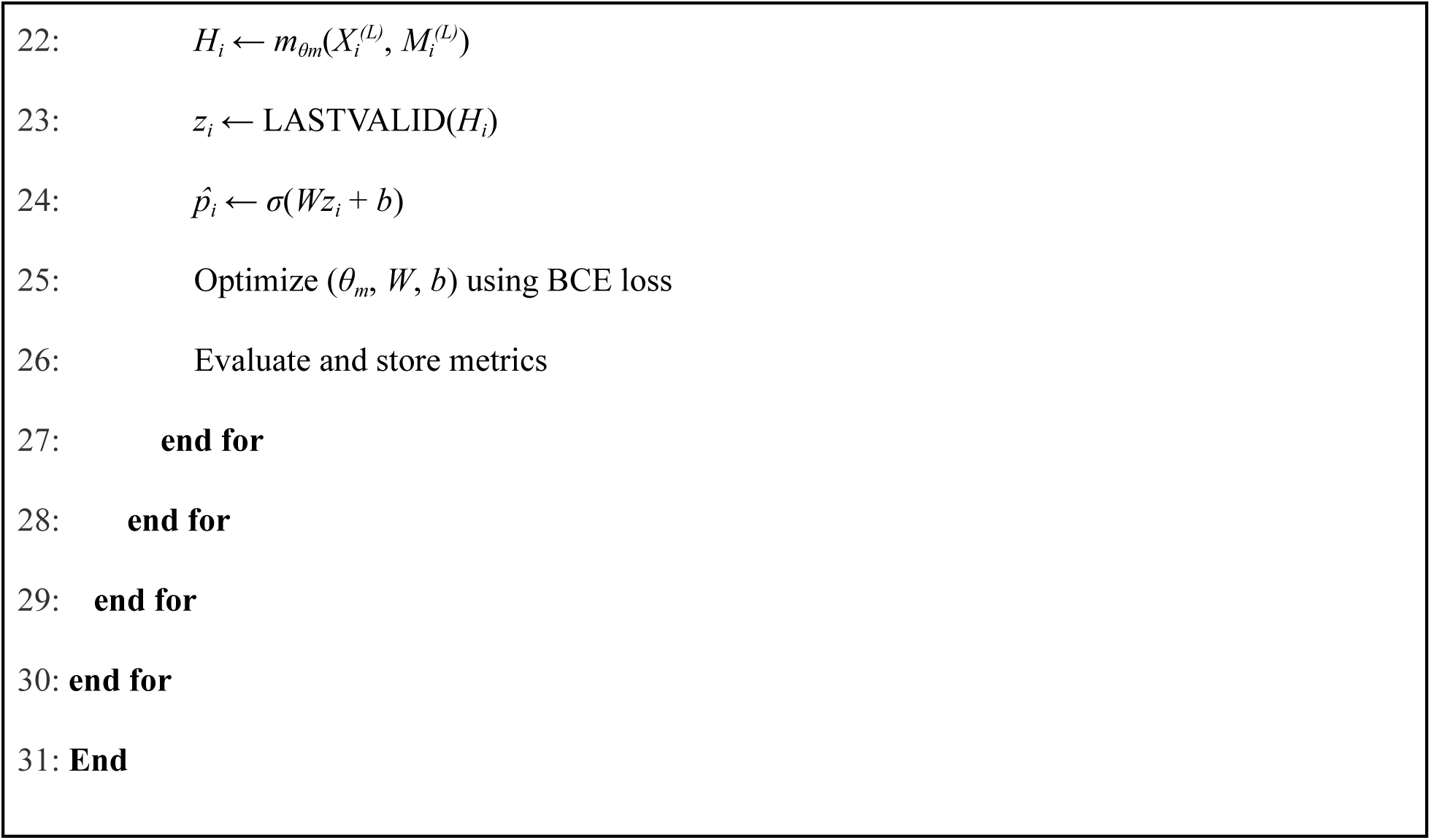

### Prognostic Model Development

#### Sequential Deep Learning Architectures

We evaluated three sequential architectures: RNN, LSTM, and Transformer encoder.^45,46^ All models were implemented in PyTorch with constrained parameterization to prevent overfitting given the limited sample size.

#### Model Specifications

- RNN: Single hidden layer (hidden size = 64), dropout regularization (p = 0.2), and fully connected output layer for binary classification
- LSTM: Identical configuration to RNN but utilizing LSTM cells
- Transformer: Single-layer encoder with one attention head and 32-dimensional embedding space.

Input features underwent learnable linear transformation with positional encodings

All temporal models were trained independently for each task, prediction horizon, feature encoder, and sequence length configuration, producing a single progression probability per patient based on the most recent valid visit, as summarized in Algorithm 2

### Training and Evaluation

For each prediction horizon, patients were labeled as progressors if disease conversion occurred within the specified time window following the index visit. Patients were labeled as non-progressors only if follow-up duration exceeded the prediction horizon without observed conversion. Otherwise, patients were excluded for that horizon to avoid mislabeling cases with insufficient follow-up (censored cases).

#### Data Splitting Strategy

All experiments employed patient-level data partitioning (80/20 split) to prevent information leakage. The NGA to GA cohort was divided into 25 training and 7 testing patients, while the NCGA to CGA cohort was split into 28 training and 7 testing patients. Stratified sampling preserved the natural distribution of progressors and non-progressors. To ensure robust performance assessment, all experimental configurations were executed across five independent random seeds, with final results reported as averaged metrics.

#### Model Optimization

Binary cross-entropy loss^45^ served as the objective function. Model parameters were optimized using the Adam optimizer^47^ with a learning rate of 1×10^−3^. Training proceeded for 10 epochs using mini-batches of 4 patients. All models were trained using a high-performance computing environment equipped with NVIDIA GeForce GTX 1080 Ti GPUs (11GB VRAM per GPU) running CUDA version 12.2 and NVIDIA driver 535.230.02.

## Results

The Wake Forest University School of Medicine cohort consisted of 91 patients contributing 455 OCT volumes acquired across multiple clinical visits. Baseline demographic and ocular characteristics are summarized in Table 1. Among 686 visits from 91 patients with valid paired grades, exact agreement between graders was 82.8%, corresponding to a Cohen’s κ of 0.70 and a quadratic-weighted κ of 0.79, indicating substantial inter-rater reliability. Discrepant cases were subsequently reviewed in a joint adjudication session to establish a final reference standard. Concordant grades were accepted as final without modification; disagreements were resolved by consensus. Agreement between Grader 1 and the adjudicated reference standard was 93.6% (κ = 0.89; weighted κ = 0.90), with 6.4% of visits modified following adjudication. Agreement between Grader 2 and the final reference standard was 89.0% (κ = 0.81; weighted κ = 0.90), with 11.0% of visits differing from the adjudicated grade. Primary model development was conducted using Grader 1 labels, with consensus adjudication serving to quantify inter-rater reliability and define the reference standard for discrepant cases.

**Table 1:**
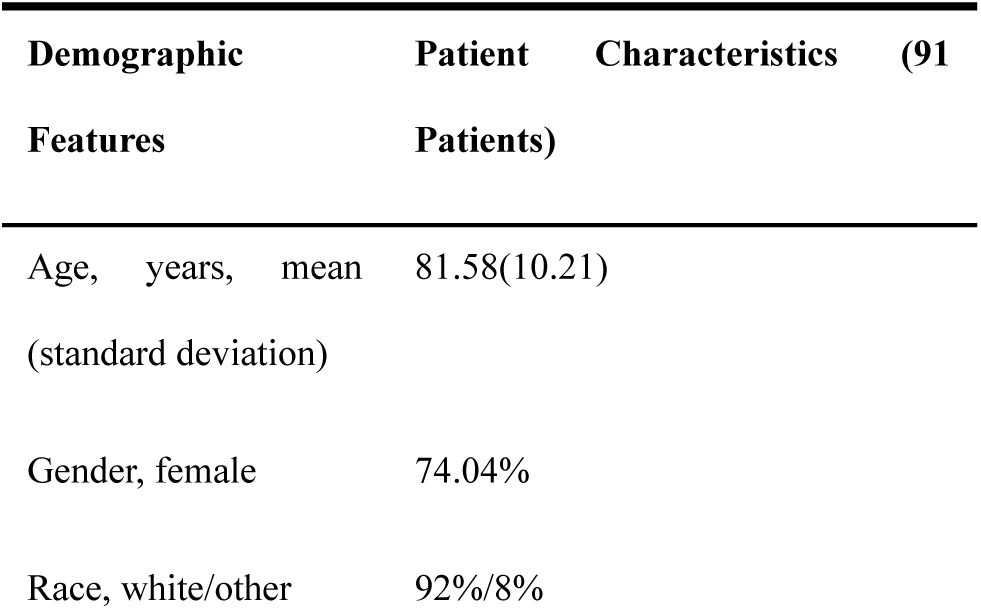
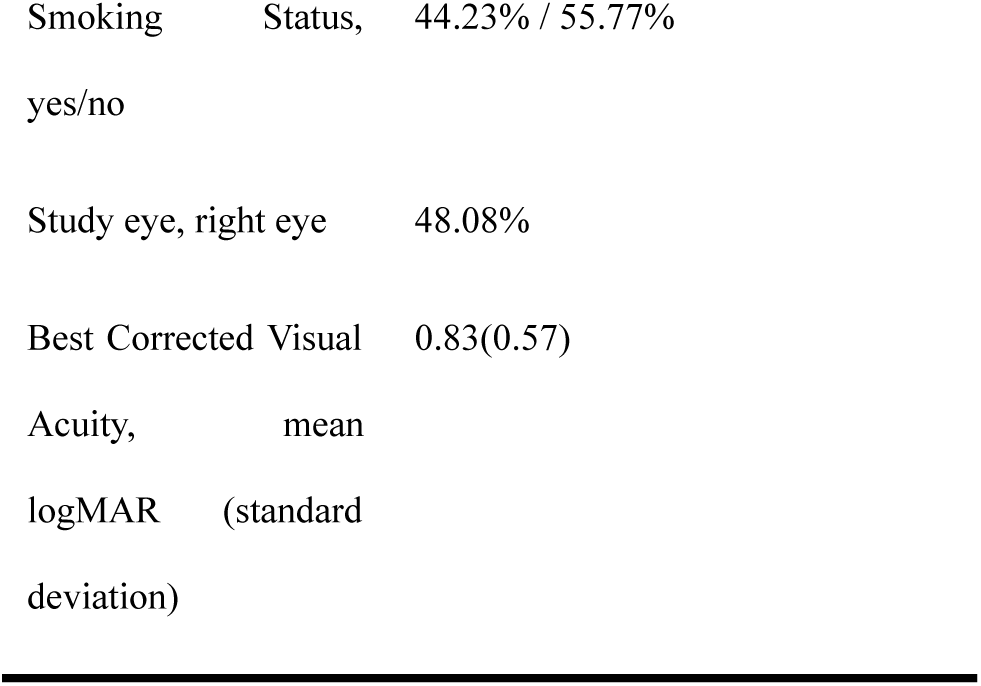
Baseline Demographic And Ocular Characteristics for Wake Forest Dataset.

Longitudinal deep learning models demonstrated consistent predictive performance for both GA progression tasks- NGA to GA and NCGA to CGA across 2-to-6-year prediction horizons. Performance was evaluated using ROC-AUC as the primary metric, with F1-score and accuracy as complementary measures. OCT visits were encoded as temporal sequences and modeled using RNN, LSTM, and Transformer architectures. Figure 4 presents the best-performing configurations for each architecture, sequence length, and horizon.

### NGA to GA Progression Prediction

Longitudinal models achieved robust performance for NGA to GA prediction across all horizons. At 2-4 years, ROC-AUC values ranged from 0.84 to 0.94 across architectures, backbones, and sequence lengths, with standard deviations generally ≤ 0.15. At 5-6 years, multiple configurations achieved ROC-AUC of 1.00 ± 0.00 with high F1-scores and accuracies, consistent across random seeds despite reduced long-term non-progressors. Both shorter (7-visit) and longer (19-visit) sequences achieved comparable performance, with no architecture uniformly dominating. NGA to GA prediction demonstrated the highest and most stable performance of both tasks, remaining substantially above chance across all horizons.

### NCGA to CGA Progression Prediction

NCGA to CGA prediction exhibited greater variability and lower performance than NGA to GA (Table 3). At 2–3 years, ROC-AUC ranged from 0.60 to 0.80 with larger standard deviations. Performance improved at 4–5 years, with Transformer models achieving peak AUC of 0.95 ± 0.11. At 6 years, AUC values ranged from 0.70 to 0.80. Longer sequences (8 visits) consistently outperformed shorter sequences (4 visits), particularly at intermediate and extended horizons. Transformers achieved the highest AUCs more frequently than RNN or LSTM models. While performance was lower and more variable than NGA to GA, longitudinal modeling enabled reliable risk stratification, remaining consistently above chance.

**Table 2:** Performance comparison across horizons, sequence lengths, and models for NGA to GA progression prediction. All values are reported as mean ± std.

| Horizon<br>(Years) | Seq. Length<br>(no of visits) | Backbone | Temporal Model | $\Delta t$ used | AUC<br>(mean ± std) | F1<br>(mean ± std) | Accuracy<br>(mean ± std) |
| --- | --- | --- | --- | --- | --- | --- | --- |
| 2 | 7 | ResNet-18 | LSTM | No | $0.940 \pm 0.089$ | $0.856 \pm 0.031$ | $0.771 \pm 0.078$ |
| | 19 | ResNet-50 | LSTM | No | $0.940 \pm 0.055$ | $0.848 \pm 0.034$ | $0.743 \pm 0.064$ |
| 3 | 7 | ResNet-50 | RNN | No | $0.880 \pm 0.110$ | $0.840 \pm 0.089$ | $0.771 \pm 0.128$ |
| | 19 | ResNet-18 | LSTM | Yes | $0.900 \pm 0.141$ | $0.800 \pm 0.075$ | $0.686 \pm 0.064$ |
| 4 | 7 | ResNet-50 | RNN | No | $0.880 \pm 0.110$ | $0.840 \pm 0.089$ | $0.771 \pm 0.128$ |
| | 19 | ResNet-18 | LSTM | Yes | $0.900 \pm 0.141$ | $0.800 \pm 0.075$ | $0.686 \pm 0.064$ |
| 5 | 7 | ResNet-50 | RNN | No | $1.000 \pm 0$ | $0.966 \pm 0.046$ | $0.943 \pm 0.078$ |
| | 19 | ResNet-18 | Transformer | No | $1.000 \pm 0$ | $0.923 \pm 0$ | $0.857 \pm 0$ |
| 6 | 7 | ResNet-50 | RNN | No | $1.000 \pm 0$ | $0.966 \pm 0.046$ | $0.943 \pm 0.078$ |
| | 19 | ResNet-50 | RNN | No | $1.000 \pm 0$ | $0.966 \pm 0.046$ | $0.943 \pm 0.078$ |

**Table 3:** Performance comparison across horizons, sequence lengths, and models for NCGA to CGA progression prediction. All values are reported as mean ± std.

| Horizon<br>(Years) | Seq. Length<br>(no of visits) | Backbone | Temporal Model | $\Delta t$ used | AUC<br>(mean ± std) | F1<br>(mean ± std) | Accuracy<br>(mean ± std) |
| --- | --- | --- | --- | --- | --- | --- | --- |
| 2 | 4 | ResNet-18 | Transformer | Yes | $0.792 \pm 0.195$ | $0.647 \pm 0.190$ | $0.629 \pm 0.163$ |
| | 8 | ResNet-18 | Transformer | Yes | $0.842 \pm 0.209$ | $0.629 \pm 0.255$ | $0.686 \pm 0.186$ |
| 3 | 4 | ViT-B/16 | Transformer | No | $0.792 \pm 0.102$ | $0.677 \pm 0.196$ | $0.600 \pm 0.119$ |
| | 8 | ViT-B/16 | Transformer | No | $0.775 \pm 0.242$ | $0.661 \pm 0.178$ | $0.657 \pm 0.128$ |
| 4 | 4 | ResNet-50 | Transformer | Yes | $0.883 \pm 0.095$ | $0.800 \pm 0.074$ | $0.714 \pm 0.143$ |
| | 8 | ResNet-50 | Transformer | Yes | $0.950 \pm 0.112$ | $0.816 \pm 0.146$ | $0.828 \pm 0.120$ |
| 5 | 4 | ViT-B/16 | Transformer | No | $0.900 \pm 0.141$ | $0.756 \pm 0.155$ | $0.657 \pm 0.163$ |
| | 8 | ViT-B/16 | Transformer | No | $0.960 \pm 0.055$ | $0.897 \pm 0.069$ | $0.828 \pm 0.120$ |
| 6 | 4 | ViT-B/16 | Transformer | No | $0.800 \pm 0.212$ | $0.838 \pm 0.032$ | $0.743 \pm 0.064$ |
| | 8 | ResNet-50 | Transformer | Yes | $0.780 \pm 0.268$ | $0.812 \pm 0.127$ | $0.714 \pm 0.175$ |

**Figure 4:**
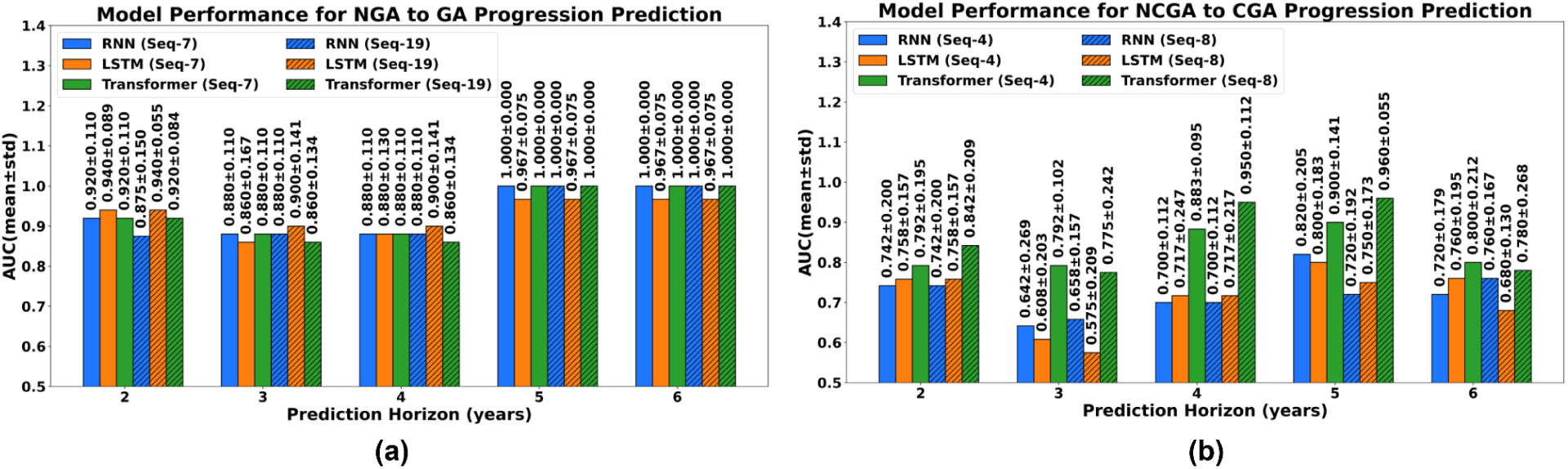
Best-performing configurations for each architecture, sequence length, and horizon. (a) NGA to GA progression prediction; (b)NCGA to CGA progression prediction.

## Discussion

This study demonstrates that longitudinal deep learning modeling of OCT sequences can predict GA progression across multiple clinically relevant prediction horizons/follow-up periods ranging from 2 to 6 years. By processing sequential visit-level embeddings through RNNs, LSTMs, and Transformers, our framework enables individualized risk prediction for both GA onset (NGA to GA) and central involvement (NCGA to CGA). Despite the limited cohort size, the models consistently achieved meaningful predictive performance with incorporation of inter-visit time intervals (Δt) improving stability in several configurations.

Performance gains stem from explicitly modeling disease dynamics rather than isolated imaging snapshots. GA progresses over time with subtle changes in the retinal pigment epithelium and outer retinal layers that may not be detected at single visits.^9,17,22^ By converting each OCT visit into a numerical representation and arranging these chronologically as a sequence, the models learn disease progression trajectories while reducing imaging noise and capturing the longitudinal relationships between disease changes across successive visits, even when visit intervals are irregular; a critical requirement for real-world clinical data. Unlike prior cross-sectional approaches focused on GA detection,^28,30^ short-term growth prediction from single baseline images^26,27^ or binary conversion with fixed follow-up intervals^29,33^ our framework uniquely models multi-year disease state transitions (NGA to GA and NCGA to CGA) using longitudinal sequences with irregular spacing, extending to 5-6 year forecasting directly relevant to long-term treatment planning. Additionally, using visit-level labels rather than pixel-level annotations substantially reduces annotation burden while enabling analysis of larger longitudinal cohorts Our dataset included patients with extended follow-up, with some patients observed for more than 10 years, compared with typical AMD studies spanning 1-3 years.^28^ This extended observation period enables assessment of longer follow-up periods critical for treatment planning but also results in progressive class imbalance reflecting the natural history of GA.

For NGA to GA prediction, progressor prevalence increased from 65.6% at 2 years to 87.5% at 5-6 years. With approximately 6-7 test patients per split, this corresponds to 4-5 progressors at the 2-year prediction horizon, enabling robust discrimination (AUC up to 0.940 ±0.089), but only 0-1 non-progressors at extended prediction horizons. While such extreme imbalance limits formal statistical interpretation, perfect discrimination at 5-6 years (AUC 1.000±0.000) demonstrates successful identification of rare slow progressors, a clinically valuable population for extended monitoring. At intermediate prediction horizons (2-4 years) with more balanced distributions, ResNet-50 combined with RNN or LSTM architectures achieved AUCs between 0.880 and 0.940, providing strong evidence of discriminative capability. NCGA to CGA conversion reached 68.6% at 6 years, consistent with published Kaplan–Meier estimates of approximately 57% at 4 years and 60% at 5 years in AREDS2.^16,48^ For this task, more balanced class distributions across all horizons (51.4-68.6% progression) enabled stable performance assessment. Transformer-based architectures demonstrated strong and consistent performance, with AUCs of 0.950±0.112 and 0.960±0.055 at 4- and 5-year horizons, respectively. Temporal interval encoding (Δt) specifically benefited LSTM-based models at intermediate horizons, indicating that explicitly modeling visit spacing helps distinguish physiological aging from pathological progression.

Analysis of sequence length revealed task-dependent temporal requirements. For NGA to GA prediction, shorter (7-visit) and longer (19-visit) sequences achieved comparable performance, suggesting that recent disease trajectory may be sufficient to predict atrophy onset. In contrast, for NCGA to CGA prediction, longer sequences (8 visits) outperformed shorter sequences (4 visits) at extended prediction horizons, implying that prediction of foveal involvement benefits from integrating longer-term spatial progression patterns. These findings suggest that different disease transitions operate on distinct temporal scales.

From a clinical perspective, accurate NCGA to CGA prediction is particularly impactful given recent FDA approval of complement inhibitors demonstrating location-dependent efficacy, with greater slowing of growth rate in extrafoveal lesions.^39,40^ Given the high burden of injections (monthly or bimonthly), there is now increasing interest in specifically targeting patients who are fast progressors towards the fovea, as they are at greater risk of significant visual impairment. Multi-horizon predictions align with real-world decision-making, where management depends on anticipated disease trajectory rather than binary outcomes. Such models can enable risk-based treatment prioritization, identifying patients at highest risk of irreversible central vision loss before foveal involvement occurs. Prospective validation comparing AI-guided surveillance against standard care will be required before clinical deployment.

This study establishes feasibility and lays groundwork for several promising hypotheses. Future work incorporating multi-rater annotations would strengthen label reliability and enable assessment of inter-observer variability. Several limitations should be acknowledged in the context of the exceptional rarity of longitudinal OCT datasets in which disease stage (NCGA or CGA) was systematically recorded at each clinic visit over time. To our knowledge, no prior study has modeled multi-year NCGA to CGA progression from sequential OCT visits, and the assembly of this decade-long dataset represents a significant contribution in itself. While our cohort size is modest (n=32-35 per prediction task), this reflects the inherent scarcity of longitudinal OCT data with granular visit-level NCGA and CGA staging rather than a methodological limitation. Our dataset compiled over a decade (2013-2023) represents one of the most comprehensively staged longitudinal GA datasets reported to date. The extended longitudinal follow-up spanning greater than 10 years is unusually comprehensive for GA research, enabling investigation of clinically relevant 5–6-year follow-up periods rarely examined in prior studies. The progressive imbalance between progressors and non-progressors at extended prediction horizons authentically reflects natural GA progression patterns, validating our cohort’s representativeness while highlighting opportunities for larger multi-center efforts. External validation across institutions, OCT platforms, and diverse populations is challenging given the field-wide scarcity of comparable longitudinal datasets with visit-level NCGA and CGA labels; establishing collaborative multi-center efforts to aggregate such rare data represents a critical priority for the field. Additionally, progression labels were defined based on anatomical OCT staging without incorporating functional outcomes such as visual acuity; however, anatomical progression, particularly foveal involvement, is strongly associated with clinically meaningful vision loss and has emerged as a key consideration in the era of complement inhibitor therapies demonstrating greater efficacy in extrafoveal lesions. Integrating multimodal imaging and clinical risk factors such as genetic profiles and imaging biomarkers represents a natural next step that could enhance predictive accuracy beyond OCT alone. The retrospective design with variable visit intervals reflects real-world clinical practice; prospective studies with standardized protocols would enable more controlled validation. As our cohort is predominantly White from an urban center in the Southeastern United States, generalizability to diverse populations remains to be established. Clinical adoption will also require improved interpretability, such as attention visualization or gradient-based attribution methods, to ensure that models rely on clinically meaningful features.

In conclusion, this work establishes that temporal deep learning can forecast GA progression across clinically meaningful multi-year prediction horizons/follow up periods using routine OCT imaging, advancing beyond predominantly short-term approaches in prior literature. Although larger multi-center validation is required, the demonstrated feasibility of modeling long-term disease trajectories provides a foundation for automated risk stratification systems. As the newly approved therapies transform GA management, such AI tools may enable precision medicine approaches that match treatment decisions to an individual’s risk of progression, ultimately preserving vision through timely intervention.

## Data Availability

All data produced in the present study are available upon reasonable request to the authors

## List of abbreviations

AMD: Age-related macular degeneration
GA: geographic atrophy
NGA: no geographic atrophy
NCGA: non-central geographic atrophy
CGA: central geographic atrophy
RPE: retinal pigment epithelium
iRORA: incomplete retinal pigment epithelium and outer retinal atrophy
cRORA: complete retinal pigment epithelium and outer retinal atrophy
OCT: optical coherence tomography
FAF: fundus autofluorescence
ETDRS: Early Treatment Diabetic Retinopathy Study
BCVA: best corrected visual acuity
LogMAR: logarithm of the minimum angle of resolution
AI: artificial intelligence
CNN: convolutional neural network
RNN: recurrent neural network
LSTM: long short-term memory
ViT: vision transformer
ViT-B/16: Vision Transformer Base/16
ResNet-18, ResNet-50: residual neural network
ROC-AUC: receiver operating characteristic area under the curve
Δt: F1-score delta time between visits
RGB: red, green, blue
FDA: U.S. Food and Drug Administration
IRB: institutional review board
ICD-9, ICD-10: International Classification of Diseases
NEI: National Eye Institute
VEGF: vascular endothelial growth factor
SPIE: International Society for Optics and Photonics
ARVO: Association for Research in Vision and Ophthalmology
OD: right eye
OS: left eye

## Declaration of generative AI in scientific writing

During the preparation of this work the authors used generative artificial intelligence tools for English language editing and grammar correction. After using this tool, the authors reviewed and edited the content as needed and took full responsibility for the content of the publication.

## Author Contributions

Sadia Siraz: Methodology, data analysis, model development, visualization, writing - original draft.

Hindolo Kamanda: Data collection.

Sina Gholami, Ahammed Sakir Nabil: Validation, writing - review & editing.

Nitya Tangada Rao, MD: Data label grading (2nd), writing - review & editing

Sally Shin Yee Ong, M.D: Data label grading (1st), Supervision, writing - review and editing

Minhaj Nur Alam, Ph.D.: Conceptualization, supervision, funding acquisition, writing - review & editing.

All authors read and approved the final manuscript.

